# Association between diabetes and levels of micro-nutrients in a Middle Eastern population - A case-control study

**DOI:** 10.1101/2023.07.11.23292494

**Authors:** Nada Soliman, Ruba Almishal, Basant Elsayed, Ayaaz Ahmed, Sara Al-Amri, Aisha Al-Kuwari, Shaikha Al-Muhannadi, Muhammed Nadeer, Tawanda Chivese

**Affiliations:** College of Medicine, QU Health, Qatar University, Doha, Qatar

**Keywords:** Diabetes mellitus, diabetes control, microelements, macro elements, vitamins, magnesium, Qatar Biobank (QBB)

## Abstract

**Objective:** To investigate associations between levels of micronutrients and diabetes, controlled and uncontrolled diabetes.

**Methods:** A case-control study, matched on age and gender, was performed on participants with (cases) and without diabetes (controls), who were Qatari or long-term residents. Participants with diabetes were divided into those with controlled and uncontrolled diabetes using an HbA1c cutoff of 7%. Levels of micronutrients were measured from serum and categorized into normal and abnormal levels.

**Results:** A total of 1118 participants (374 cases and 744 controls) were included with a mean age of 41.7 years (SD 9.9), of whom 53.9% were female. Of those with diabetes, 229 had controlled diabetes and 145 had uncontrolled diabetes. Compared to those without diabetes, participants with diabetes had significantly lower mean magnesium (0.80 mmol/L (SD 0.07) vs 0.84 mmol/L (SD 0.06), respectively, p<0.001). Lower magnesium and iron were observed in participants with uncontrolled compared to participants with controlled diabetes. After multivariable logistic regression, diabetes was associated with hypomagnesemia (OR 3.2, 95% CI 3.4-213.9), and low iron (OR 1.49, 95% CI 1.03-2.15). Uncontrolled diabetes showed stronger odds of association with hypomagnesemia (OR 5.57, 95% CI 3.65-8.52).

**Conclusion:** In an affluent setting in the Middle East and North Africa (MENA) region, diabetes was associated with low magnesium and low iron, and this association was worse in individuals with uncontrolled diabetes.

## Introduction

The worldwide prevalence of diabetes mellitus is increasing and estimated to rise from 463 million in 2019 to 700 million by 2045 (1). The Middle East and North Africa (MENA) region has one of the highest age-adjusted comparative prevalence of diabetes in people aged 20–79 years at 12.2% (1). With a prevalence of diabetes around 16%, Qatar has the highest expenditure per person at approximately $1,751 for diabetes (1). Uncontrolled diabetes is associated with a higher risk of complications and the need for frequent hospitalizations, which are associated with a higher cost of diabetes care. Therefore, there is a need to explore factors that may affect both the development and better control of diabetes in the MENA region.

Alterations in micronutrient intake and serum levels have been implicated in both the development and control of diabetes. Micronutrients are nutrients that the body requires in small amounts and can be divided into macro-elements, micro-elements (trace elements), and vitamins. These elements are involved in processes related to glucose metabolism; thus, they have been targeted as possible preventive or management options for diabetes. Macro-elements, such as calcium (Ca), magnesium (Mg), and iron (Fe), are elements required by adults in amounts exceeding 100 mg/day, while micro-elements like zinc (Zn) and copper (Cu) are needed in amounts between 1-100 mg/day (2, 3), Mg is involved in glucose metabolism (4), too much Fe can cause oxidative damage to β-cells, and Zn is crucial in insulin production (5). Vitamins also contribute to many different biological processes and have attracted some interest either as preventive or therapeutic agents for diabetes (6-8). Vitamin B9 (folate) and vitamin B12 aid in the conversion of homocysteine to methionine, thus reducing oxidative damage, which is common in diabetes, and frequently associated with peripheral neuropathy (9).

Despite the possible links between micro-nutrients and glucose metabolism, findings from both observational and experimental studies about their role in the development and control of diabetes have largely been heterogeneous. Several meta-analyses of RCTs (10-12) have produced conflicting results about the benefits of supplementation with micronutrients in individuals with diabetes, with some showing benefit and others showing no benefit. Therefore, there is a need to investigate the relationship between the levels of micronutrient levels and diabetes as well as whether these micronutrients play a part in the control of diabetes. In the MENA region, only a few studies have investigated the relationship between diabetes and the levels of micronutrients, and studies are needed in this region which has a unique a high diabetes prevalence. The study aimed compare the levels of micro, and macro-elements, between individuals with and without diabetes. We also investigated the association between these micronutrients and uncontrolled diabetes.

## Methods

### Design, population, and setting

A case control study, using data collected by Qatar Biobank (QBB), was conducted on Qatari citizens and long-term residents. The QBB is a population-based research initiative that creates a depository of biological samples and information on the health and lifestyle of Qatari citizens and long-term residents, aiming to prospectively recruit 60,000 volunteers (13, 14). Participants above 18 years of age with data on micronutrients were eligible for inclusion in the present study. During this study, participants with diabetes were randomly selected from the QBB database and matched, using age, gender and nationality, to those without diabetes. Participants without data on micronutrients, those aged below 18 years and pregnant women were excluded.

### Data collection

Each participant completed a questionnaire, had anthropometry measured, and had blood drawn for measuring serum micronutrients. The data collected for each participant were: demographics, data on self-reported doctor-diagnosed diabetes, duration of diabetes, nutritional supplement usage, income, smoking status, blood pressure, anthropometry, HbA1c level, serum levels of lipids, random glucose, and micronutrients. The micronutrients measured were copper (Cu), zinc (Zn), magnesium (Mg), iron (Fe), folate, and vitamin B12.

### Diabetes measurements

Participants were classified into the diabetes group if they self-reported being diagnosed by a doctor or if they had an HbA1c ≥6.5% or a random blood glucose level ≥11.1 mmol/L, according to the American Diabetes Association (ADA) criteria (14). Controlled diabetes was defined based on ADA guidelines as HbA1c ≤ 7% (13).

### Micro, macro elements and vitamins measurements

For each participant, micronutrients were assessed from serum. Levels of micronutrients were classified as ‘low’ according to the normal laboratory ranges for adults on UpToDate (14), using the following cut-offs: Mg (≤0.66mmol/L), Fe (≤9 μmol/L), Zn (≤11.5μmol/L), Cu (≤15.7μmol/L), folate (≤4.1nmol/L), and vitamin B12 (≤147.5pmol/L).

### Ethics and informed consent

At the QBB, participants gave written informed consent, before data were collected. This study received ethical approval from the Qatar University’s Institutional Review Board (QU-IRB) (reference number: QU-IRB1228-E/20), and the Qatar Biobank’s IRB (reference number: QF-QBB-RES-ACC-0186).

### Data analysis

Histograms were used to check for the normality of data distribution for measured variables. Descriptive data were presented as mean and standard deviation (SD) if normally distributed, or as median and interquartile range (IQR) if not normally distributed. Categorical variables were presented using frequencies and percentages. Boxplots were used to compare the levels of the micronutrients between participants without diabetes, those with controlled and participants with uncontrolled diabetes. The micronutrient levels were compared between participants with and without diabetes using the t-test for independent groups if normally distributed or Wilcoxon rank-sum test if not normally distributed. Comparisons of micronutrient levels between the three groups (controlled, uncontrolled, and no-diabetes) were tested using one-way ANOVA if data were normally distributed, or Dunntest if data were not normally distributed with post hoc test done using Bonferroni adjustment.

Multivariable logistic regression was carried out for the association between diabetes and the micronutrients, with separate models for each micronutrient. To investigate the association between micronutrients and uncontrolled diabetes, multivariable multinomial logistic regression was used, with “no diabetes” as the base-outcome. In all models, we adjusted for age, gender, BMI, smoking, and income, as they are known to be associated with both micronutrients’ intake and diabetes (15-17). For each multivariable regression model, we tested model specification using linktest. Exact p-values were reported and interpreted as evidence against null hypothesis and 95% confidence intervals (95% CI) were reported for odds ratios (OR). All data analyses were performed using Stata 16.0 statistical software.

## Results

### Characteristics of participants by diabetes status

A total of 1118 participants, of which 33.5% had diabetes, were included in the study. Participants with diabetes were slightly older than those without (mean age 44.4 years old (9.4) vs 40.3 years old (9.9), P<0.001). Participants with diabetes compared to those without, had significantly higher mean BMI (31.5 kg/m^2^ (SD 5.4) vs 29.8 kg/m^2^ (SD 5.6), respectively, P<0.001) (Table 1).

**Table 1.**
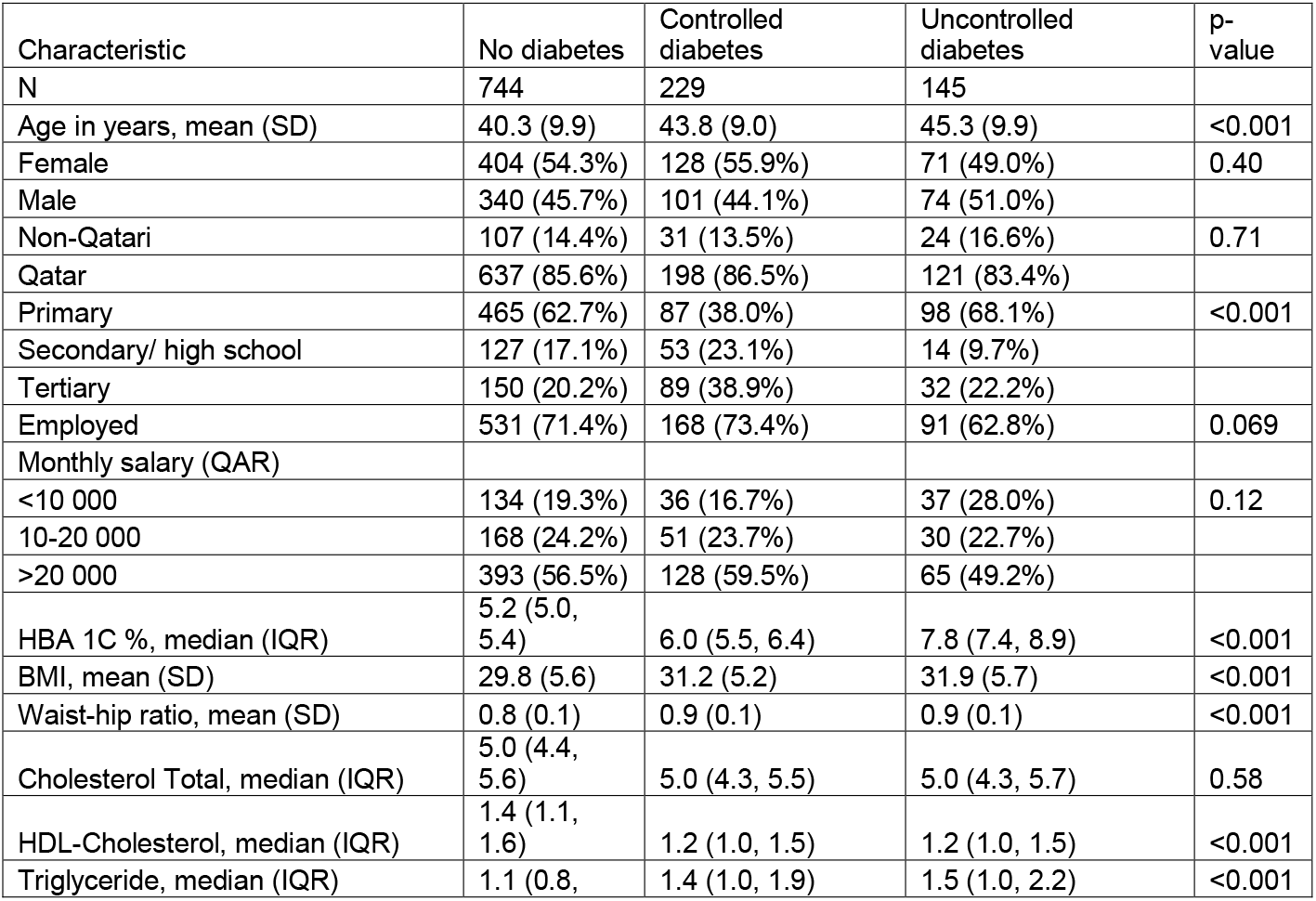
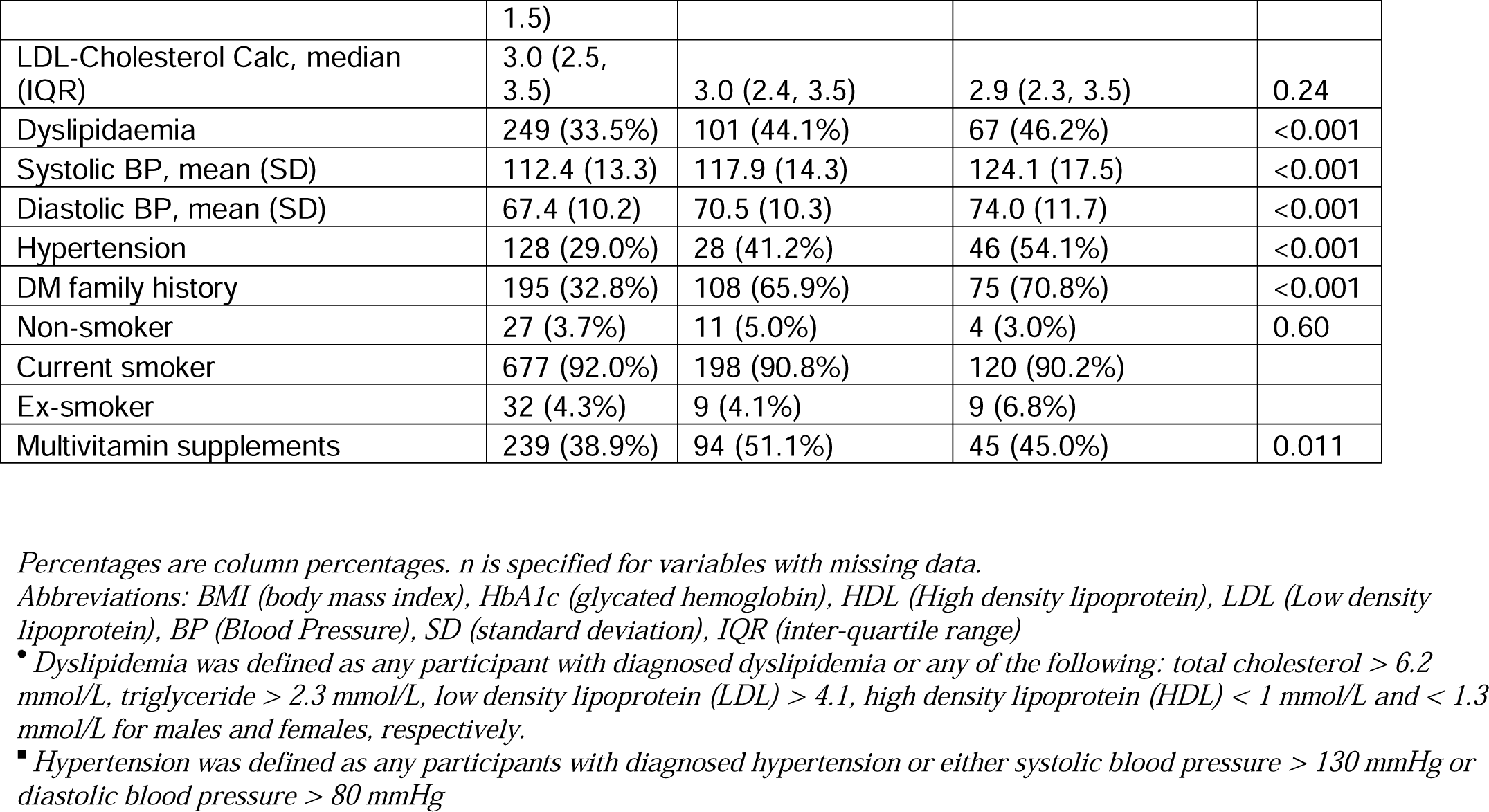
Characteristics of participants by diabetes status

### Comparison of characteristics by controlled diabetes status

As expected, median HbA1C was higher in the participants with uncontrolled diabetes, compared to those with controlled diabetes (7.8% (IQR 7.4-8.9) vs 6.0% (IQR 5.5-6.4), respectively, P<0.001) (Table 1). The median duration of diabetes for participants with uncontrolled diabetes was longer compared to the participants with controlled diabetes (7 years (IQR 4-13) vs 4 years (IQR 2-9), respectively, P<0.001). A greater percentage of participants with controlled diabetes reported taking multi-vitamins/minerals supplements compared to those with uncontrolled diabetes (51.1% vs 45.0%, respectively, P=0.011)

### Comparison of micronutrients levels between participants with and without diabetes

Median serum levels of iron and folate were significantly higher in participants with diabetes compared to those without (Table 2). Median Mg serum levels were significantly lower in participants with diabetes, compared to those without diabetes (0.79 mmol/L (SD 0.07) vs. 0.84 mmol/L (SD 0.06), P<0.001). For categorized micronutrients, compared to participants without diabetes, a significantly higher proportion of participants with diabetes had low magnesium (48.9% vs 22.3%, p<0.001) (Table 2).

**Table 2.**
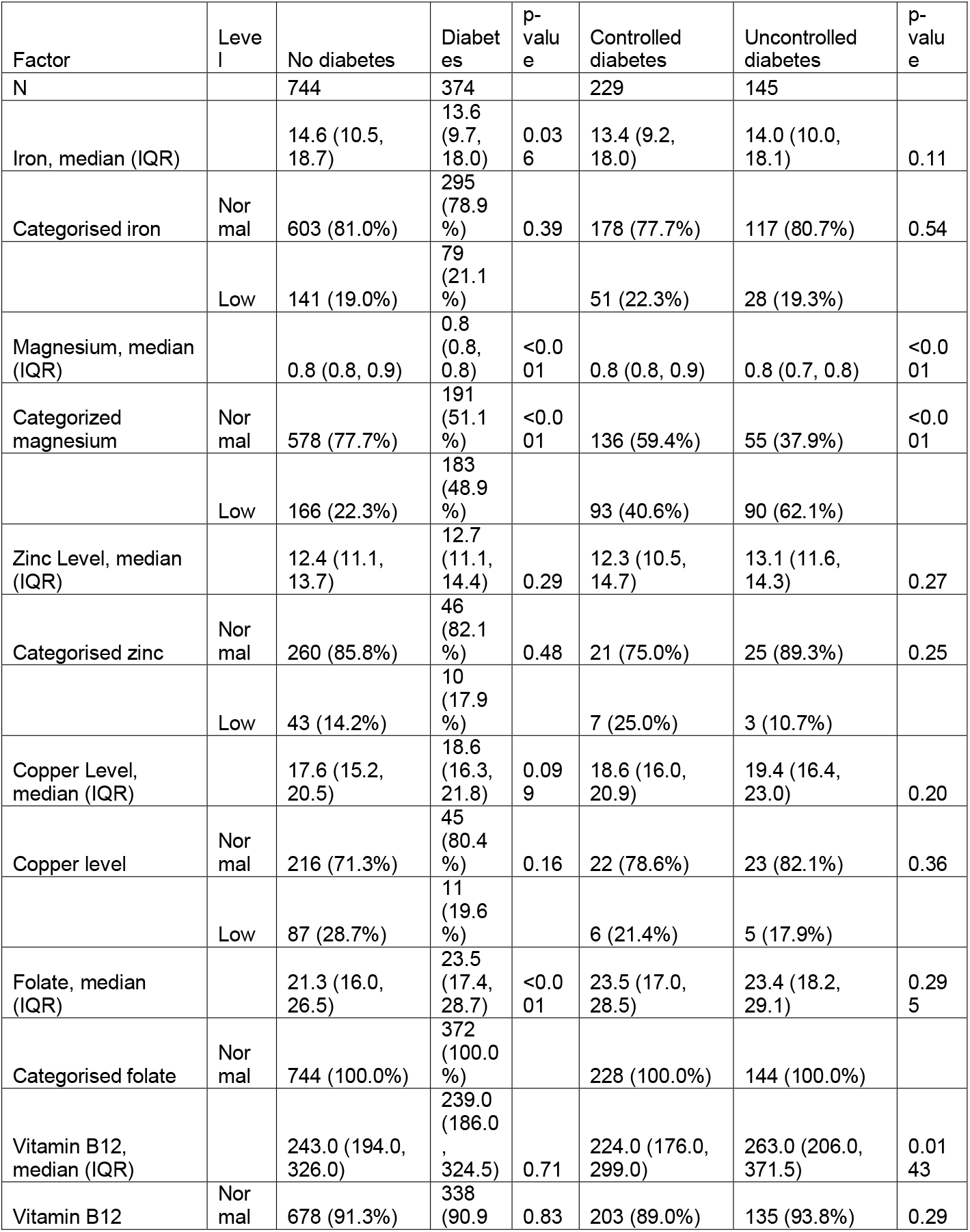
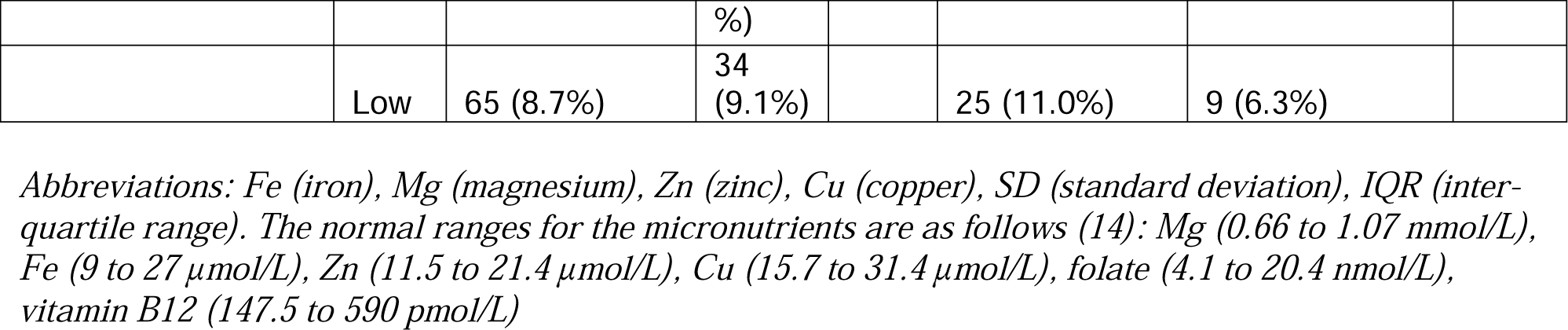
Comparison of nutrients levels between participants with and without diabetes

The boxplots in Figure 1 and Table 2 show a comparison of the levels of micronutrients between participants without diabetes, those with controlled diabetes and participants with uncontrolled diabetes. The mean Mg level was significantly higher in participants with controlled diabetes compared to those with uncontrolled diabetes (0.81 mmol/L (SD 0.07) vs 0.76 mmol/L (SD 0.07), respectively, P<0.001)) (Table 2 & Fig. 1). The median vitamin B12 levels were significantly higher in participants with uncontrolled diabetes compared to participants with either controlled or without diabetes (Table 2). There were no significant differences between the three groups in the remaining micronutrients (Table 2).

**Fig. 1:**
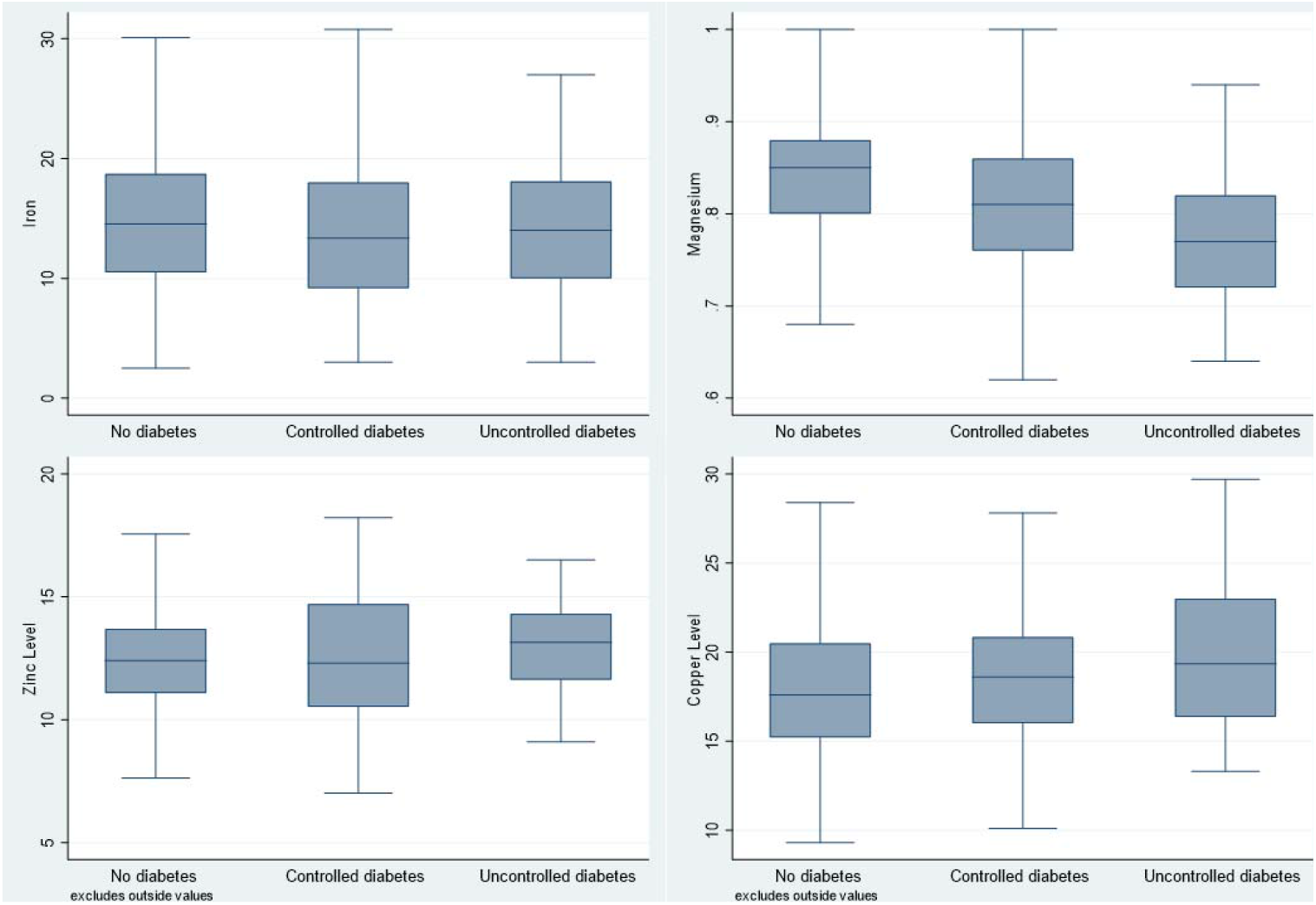
Boxplots comparing selected micronutrent levels across the three groups.

### Association between diabetes and micronutrients– multivariable logistic regression

After multivariable logistic regression, diabetes, compared to no diabetes, was significantly associated with a 3-fold odds of having low magnesium (OR 3.32, 95% CI 2.47-4.47, p<0.001), a 49% increase in the odds of having low iron (OR 1.49, 95% CI 1.03 – 2.16, p = 0.034). Diabetes was also associated with a 49% increase in the odds of having low Zn, although with weak evidence against the null hypothesis (p=0.334) (Table 3).

**Table 3.** Association between controlled diabetes and micronutrients – Multivariable multinomial logistic regression.

| Micronutrient | All Diabetes |  |  | Controlled diabetes |  |  | Uncontrolled diabetes |  |  |
| --- | --- | --- | --- | --- | --- | --- | --- | --- | --- |
|  | OR | p-value | 95%CI | OR | p-value | 95%CI | OR | p-value | 95%CI |
| Mg | 3.32 | 0.000 | 2.47 - 4.47 | 2.47 | 0.000 | 1.75 - 3.48 | 5.57 | 0.000 | 3.65 - 8.52 |
| Fe | 1.49 | 0.034 | 1.03 - 2.16 | 1.38 | 0.141 | 0.90 - 2.11 | 1.75 | 0.047 | 1.01 - 3.03 |
| Zn | 1.49 | 0.336 | 0.66 - 3.34 | 2.47 | 0.063 | 0.95 - 6.41 | 0.62 | 0.531 | 0.14 - 2.77 |
| Cu | 0.68 | 0.382 | 0.28 - 1.62 | 1.17 | 0.784 | 0.38 - 3.59 | 0.35 | 0.125 | 0.09 - 1.34 |
| Vitamin B12 | 1.17 | 0.508 | 0.74 - 1.84 | 1.37 | 0.231 | 0.82 - 2.27 | 0.84 | 0.641 | 0.40 - 1.76 |

*Abbreviations: Fe (iron), Mg (magnesium), Zn (zinc), Cu (copper), OR (odds ratio), CI (confidence interval). Adjusted for age, gender, BMI, smoking, and income.*

### Association between micronutrients and controlled diabetes – multivariable multinomial logistic regression

In further stratified analyses, uncontrolled diabetes was associated with 5.6-fold increased odds of having low magnesium (OR 5.57, 95% CI 3.65-8.57, p<0.001) and two-fold increased odds of having low iron (OR 1.75, 95% CI 1.01-3.03, p=0.047). These associations were still observed in participants with controlled diabetes, but they were noticeably weaker (Table 3). Lastly, in participants with controlled diabetes, diabetes was associated with two-and-a-half-fold increase in the odds of having low Zn, although there was weak evidence against the null hypothesis at the study’s sample size (Table 3).

## Discussion

In this case control study, we found lower serum levels for Mg and Fe in participants with diabetes, compared to those without diabetes. After adjusting for confounders, low magnesium and low iron were significantly associated with a diabetes. These association showed higher effect magnitudes in participants with uncontrolled diabetes compared to those with controlled diabetes.

Low magnesemium was significantly associated with a 3-fold increase in diabetes risk and a 6-fold increase in the risk of uncontrolled diabetes. Consistent with our finding, one study showed Mg levels were lower in uncontrolled diabetes compared to controlled diabetes (18). Further, similar to our findings, a meta-analysis of 13 cohort studies with 536,318 participants reported a significant inverse association between Mg intake and risk of type 2 diabetes (19). Experimental studies suggest that magnesium supplementation may have benefit in people with diabetes. A meta-analysis of 21 RCTs showed that supplementation with Mg significantly improved insulin sensitivity and glucose tolerance (20). Mg works as an insulin sensitizer by regulating the tyrosine kinase activity of insulin’s receptor (21). In diabetes, insulin resistance decreases renal Mg reabsorption, resulting in urinary Mg wasting. Thus, people with diabetes may end up in a vicious circle in which hypomagnesemia enhances insulin resistance and insulin resistance causes hypomagnesemia (22). Our findings, in addition to published data, suggest that magnesium supplementation may help in reducing the risk of diabetes and enhancing diabetes control.

Diabetes was associated with a 13 folds increase in the odds of having low iron, and the effect was worse in participants with uncontrolled diabetes. Further, low Zn levels were associated with diabetes although with weak evidence against the null hypothesis. Several *in vitro* (23-25) and *in vivo* studies (26) suggest that Zn may have benefits in both reducing the risk of diabetes and better diabetes control through its action on both β cell function and insulin action. Experimental studies also suggested that Zn is beneficial in better control of diabetes (27).

Our study is a case control study, but the measurements were done in a cross-sectional method, therefore, does not enable us to establish temporality, i.e. that abnormalities in the nutrients were present before the development of diabetes. Another limitation is that we were not able to collect data on physical exercise, diet, and alcohol consumption, which are important as they could be confounders. Our study is the first on this topic in Qatar, a country that has a relatively high prevalence of diabetes that is expected to increase in the future. This study provides data about the association between micronutrients and diabetes in a relatively affluent population. Our study also provides data on differences in the levels of these micronutrients between participants with controlled and uncontrolled diabetes. Whether these differences are a result of diabetes pathophysiological processes or due to nutritional supplementation, it requires further and more robust research. Another area of further research would be whether Mg supplementation may result in better levels of diabetes control, as our observational data suggest.

## Conclusion

In an affluent setting in the MENA region, diabetes was associated with low magnesium and low iron, and this association was worse in individuals with uncontrolled diabetes.

## Data Availability

All data produced in the present study are available upon reasonable request to the authors

## Abbreviations

Fe: (Iron)
Mg: (Magnesium)
Zn: (Zinc)
Cu: (Copper)
MENA: (Middle East and North Africa)
HbA1c: (glycated hemoglobin)
BMI: (Body Mass Index)
HDL: (High Density Lipoprotein)
LDL: (Low Density Lipoprotein)
BP: (Blood Pressure)
OR: (Odds Ratio)
CI: (Confidence Interval)
β: (beta coefficient)
RCT: (Randomized Control Trial)

## Acknowledgements

We would like to express our gratitude to Qatar Biobank for providing us with the needed data in a short period of time. We also would like to thank Professor Suhail Doi and Dr. George Hindy for providing guidance and useful critiques to our research work.

## Author contributions

All the authors contributed to the conception, design, conduct, analysis and writing up of the study.

